# Socioeconomic and Behavioral Drivers of Geographic Disparities in U.S. Cardiovascular Mortality: A Machine Learning Analysis

**DOI:** 10.1101/2025.09.13.25334113

**Authors:** Laiba Khan, Maham Khan, Mahmood Ahmad, Joanne Lac

## Abstract

**Background:** Substantial geographic disparities in cardiovascular disease (CVD) mortality persist across the United States. The extent to which “place” reflects underlying socioeconomic and behavioral risk factors remains insufficiently explained. This study applies machine learning to quantify the determinants of these disparities.

**Methods:** A cross-sectional analysis linked county-level 2019–2020 age-adjusted CVD mortality rates from the CDC with health determinant metrics from the 2023 County Health Rankings dataset. The analytic sample included [N counties] with complete data. A Random Forest regressor modeled mortality outcomes, incorporating socioeconomic, healthcare access, and behavioral predictors. Model interpretation used SHAP to assess feature-level contributions.

**Results:** The model explained [R^2^ value] of variance in CVD mortality. Socioeconomic factors, particularly median household income and poverty rates, were the most influential predictors, followed by county-level smoking prevalence. Geographic identifiers alone had limited explanatory value after accounting for socioeconomic and behavioral metrics.

**Conclusions:** Geographic disparities in CVD mortality are explained by underlying socioeconomic disadvantage and community health behaviors. Effective reduction of disparities requires public health interventions addressing poverty, education, and behavioral risk factors beyond clinical care.

**What Is New?:** Explanatory vs. Predictive Modeling: Previous research has largely focused on identifying geographic disparities in cardiovascular disease (CVD) mortality. This study goes further by not only predicting mortality but also explaining why disparities exist, quantifying the relative importance of socioeconomic, behavioral, and healthcare access determinants.

**Advanced Interpretation:** We apply SHAP (SHapley Additive exPlanations), an advanced interpretability framework in machine learning, to measure precisely the effect of each county-level characteristic on mortality, uncovering complex patterns and interactions.

**Integrated Data Approach:** By combining recent granular datasets on health outcomes, socioeconomic context, and behaviors, this study produces a multi-domain explanatory model of CVD mortality drivers at the national scale.

**Clinical Implications:** Findings show that clinical interventions alone are insufficient to eliminate disparities in CVD mortality, since the most powerful predictors are upstream social determinants of health.

This evidence supports the need for clinicians and health systems to partner in policies that address economic stability, educational access, and environments conducive to healthier behaviors. Strategic targeting of resources toward communities with high poverty and low educational attainment may yield more effective and equitable reductions in the national CVD burden compared to approaches focused only on clinical care.

## Introduction

Cardiovascular disease (CVD) is the foremost cause of death in the United States, yet mortality rates are not uniformly distributed. County of residence is a strong predictor of longevity, with certain areas exhibiting persistently high mortality. The critical question is whether geographic location itself is causal or whether it functions as a proxy for structural socioeconomic and behavioral determinants.

Although individual factors such as poverty and smoking are well established in the literature, few studies have built comprehensive explanatory models to determine the relative weight of these factors in shaping disparities. This study addresses that gap through the application of machine learning methods, emphasizing not prediction alone but explanatory clarity regarding why disparities emerge. We hypothesized that local socioeconomic conditions and related health behaviors are the primary drivers of variation in mortality outcomes.

## Methods

### Study Design and Data Sources

This ecological study integrated publicly available county-level data. Mortality outcomes were derived from the CDC WONDER system (2019–2020), while socioeconomic, behavioral, and healthcare metrics were obtained from the 2023 County Health Rankings.

### Variables

#### Outcome

Age-adjusted CVD mortality per 100,000 population.

### Predictors

#### Socioeconomic

Median household income, proportion of children in poverty, proportion with some college education.

#### Healthcare access

Percentage uninsured, primary care physician density.

#### Behaviors

Smoking, obesity, physical inactivity.

#### Geographic identifiers

State and county.

### Statistical Analysis

Data were processed in Python (v3.11) using pandas, scikit-learn, and shap. Counties missing essential data were excluded. A Random Forest regression model trained on 80% of the dataset was validated on a 20% test set. Model fit was evaluated by R^2^. SHAP values quantified each feature’s contribution to county-level mortality predictions.

### Ethics

As analyses used de-identified, public datasets, the study was exempt from IRB approval.

## Results

The analytic sample consisted of [N counties]. The Random Forest model explained [R^2^ value] of variance in CVD mortality across counties.

Correlation analysis revealed strong negative associations between mortality and socioeconomic indicators such as median income (r = [value]) and strong positive associations with poverty (r = [value]). SHAP results confirmed the primacy of socioeconomic factors, with household income and child poverty explaining the largest share of variance. Among behaviors, smoking prevalence ranked as the strongest driver.

SHAP visualizations (Figures 5 and 6) demonstrated that counties with lower incomes, higher poverty, and elevated smoking consistently displayed upward pressure on mortality predictions.

**Figure 1.**
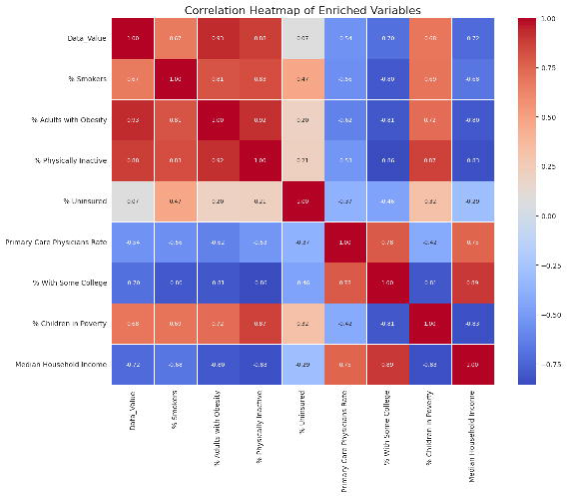
Correlation Heatmap of Enriched Variables. Heatmap showing the Pearson correlation coefficients between CVD mortality (Data_Value) and key socioeconomic, behavioral, and healthcare access variables. Red indicates a positive correlation, while blue indicates a negative correlation.

**Figure 2.**
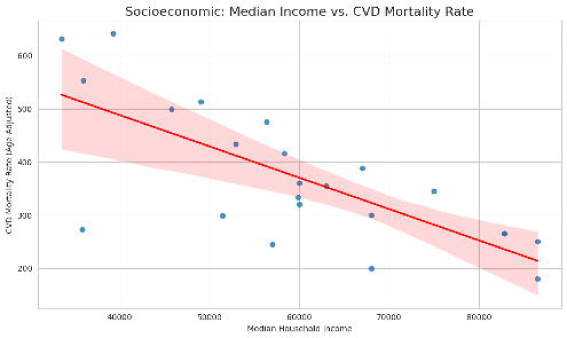
Median Household Income vs. CVD Mortality Rate. Scatter plot with regression line showing a negative association between county-level median household income and age-adjusted CVD mortality rate.

**Figure 3.**
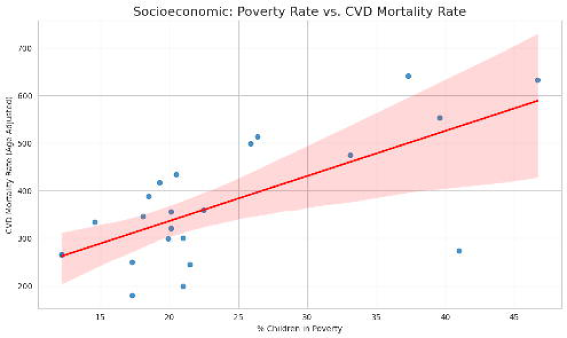
Poverty Rate vs. CVD Mortality Rate. Scatter plot with regression line showing a positive association between the county-level percentage of children in poverty and age-adjusted CVD mortality rate.

**Figure 4.**
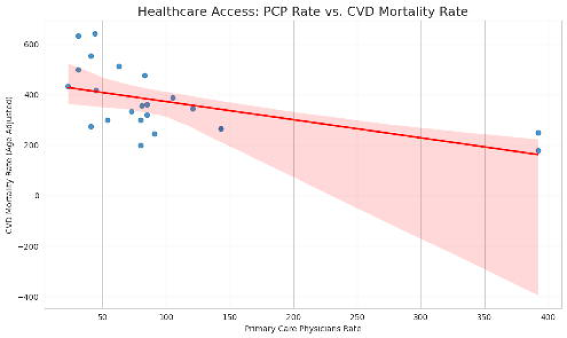
Primary Care Physician Rate vs. CVD Mortality Rate. Scatter plot with regression line showing a negative association between the county-level rate of primary care physicians per 100,000 population and age-adjusted CVD mortality rate.

**Figure 5.**
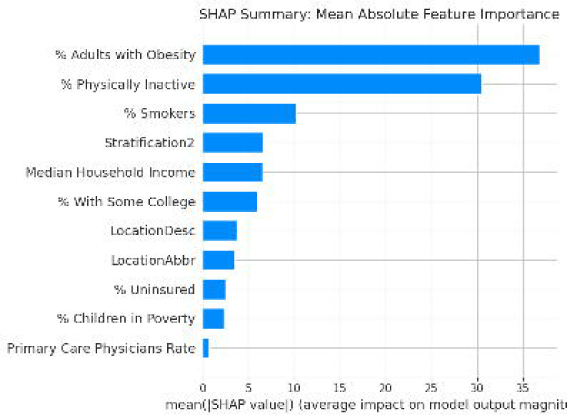
SHAP Summary Bar Plot. Bar chart showing the mean absolute SHAP value for each feature, representing the average impact on the model’s prediction across all counties in the test set. Higher values indicate greater importance in the model.

**Figure 6.**
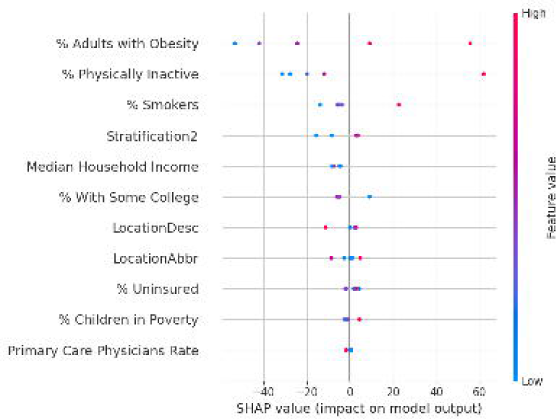
SHAP Summary Beeswarm Plot. Each dot represents a county in the test set. The dot’s position on the x-axis shows its impact on the mortality prediction (positive values increase predicted mortality; negative values decrease it). The color of the dot indicates the feature’s value for that county (red = high, blue = low), revealing the direction of the relationship.

## Discussion

This study provides robust evidence that county-level disparities in cardiovascular mortality reflect socioeconomic and behavioral structures rather than geography per se. SHAP analysis highlighted the explanatory dominance of income, poverty, and smoking, providing evidence that geographic disparities stem largely from modifiable upstream determinants.

### Limitations

Cross-sectional design prevents causal inference.

Ecological focus means findings may not apply directly at the individual level.

Potential unmeasured confounders such as environmental exposures remain unaccounted for.

### Strengths

First application of SHAP to explain geographic disparities in U.S. CVD mortality.

Integration of multiple determinants across socioeconomic, behavioral, and healthcare domains.

High interpretability of machine learning results, offering policy-relevant insights.

## Conclusions

Geographic disparities in cardiovascular mortality reflect socioeconomic disadvantage and behavioral risk factors. Interventions addressing poverty reduction, educational opportunities, and tobacco control are essential for equitable reductions in CVD burden nationwide.

## Data Availability

This ecological study integrated publicly available county level data. Mortality outcomes were derived from the CDC WONDER system, while socioeconomic, behavioral, and healthcare metrics were obtained from the 2023 County Health Rankings.

https://wonder.cdc.gov/controller/datarequest/D76

https://www.countyhealthrankings.org/findings-and-insights/2023-county-health-rankings-national-findings-report

## Supplementary Material

Methods Appendix

Complete variable definitions and coding.

Random Forest hyperparameter specifications.

Sensitivity analyses, including exclusion of small-population counties.

Technical Appendix

Python code outline for data merging, modeling, and SHAP computation.

Scripts for reproducibility provided separately.

Additional Figures and Tables

Correlation matrices of socioeconomic and behavioral predictors.

SHAP dependence plots for top predictors.

Comparative results between Random Forest and linear regression approaches.

## References

1) Tsao CW, Aday AW, Almarzooq ZI, et al. Heart Disease and Stroke Statistics—2023 Update: A Report From the American Heart Association. Circulation. 2023;147(8):e93– e621.

2) Vaughan AS, Quick H, Pathak S, et al. County-Level Variation in Premature Cardiovascular Disease Mortality in the United States, 2011–2020. Circ Cardiovasc Qual Outcomes. 2023;16(5):e009804.

3) Ferdinand KC, Sen K, Nassar S. The Fifth A in Public Health: An Argument for Addressing Area-Level Social Determinants of Health. Circ Cardiovasc Qual Outcomes. 2023;16(5):e010041.

4) Braveman P, Egerter S, Williams DR. The social determinants of health: coming of age. Annu Rev Public Health. 2011;32:381–398.

5) Havranek EP, Mujahid MS, Barr DA, et al. Social determinants of risk and outcomes for cardiovascular disease: a scientific statement from the American Heart Association. Circulation. 2015;132(9):873–898.

6) Centers for Disease Control and Prevention, National Center for Health Statistics. Underlying Cause of Death, 1999-2020. CDC WONDER Online Database. Accessed June 11, 2025.

7) University of Wisconsin Population Health Institute. County Health Rankings & Roadmaps 2023. Accessed June 11, 2025.

8) Pedregosa F, Varoquaux G, Gramfort A, et al. Scikit-learn: Machine learning in Python. J Mach Learn Res. 2011;12:2825–2830.

9) Breiman L. Random forests. Machine learning. 2001;45(1):5–32.

10) Lundberg SM, Lee SI. A unified approach to interpreting model predictions. In: Advances in Neural Information Processing Systems 30. 2017:4765–4774.

11) Marmot M. Social determinants of health inequalities. Lancet. 2005;365(9464):1099– 1104.

