## Supplementary information for "Socioeconomic and Behavioral Drivers of Geographic Disparities in U.S. Cardiovascular Mortality: A Machine Learning Analysis"

### Methods Appendix

#### Data Sources

- Mortality outcomes: Age-adjusted cardiovascular disease (CVD) mortality per 100,000 population for U.S. counties, obtained from the CDC WONDER system (2019–2020).
- Health determinants: 2023 County Health Rankings dataset, including socioeconomic, healthcare access, and behavioral measures.

#### Variables

- Outcome: Age-adjusted CVD mortality rate.
- Socioeconomic: Median household income, % children in poverty, % adults with some college.
- Healthcare access: % uninsured, primary care physician rate.
- Behaviors: % smokers, % adults with obesity, % physically inactive.
- Geographic identifiers: State, county.

#### Data Preparation

- FIPS codes were cleaned and standardized for reliable merging of datasets.
- All numeric variables were coerced to numeric type; rows with missing or invalid values were excluded.
- The merged analytic dataset included N = [final number of counties] with complete data.

#### Statistical Analysis

- Modeling approach: Random Forest regression.
- Training/validation: 80% of counties were used for model training, 20% held out for validation.
- Performance metric: R².
- Interpretability: SHAP (SHapley Additive exPlanations) was applied to quantify the marginal contribution of each predictor to model output.
- Hypothesis-driven visualizations: scatterplots of income, poverty, and physician supply against mortality; correlation heatmaps; SHAP bar and beeswarm plots.

#### Hyperparameter Specifications

- Estimator: Random Forest Regressor
- n_estimators = 100
- random_state = 42
- n_jobs = –1 (parallel processing enabled)

#### Sensitivity Analyses

- Exclusion of small-population counties with unstable mortality rates.
- Comparison of Random Forest with linear regression to test robustness of variable importance rankings.

#### Technical Reproducibility

- All analyses were conducted in Python 3.11 using:
- • pandas (data processing)
- • scikit-learn (modeling)
- • shap (interpretability)
- • matplotlib/seaborn (visualization)
- A full code outline for data cleaning, merging, model training, and SHAP computation is available upon request.
